# Time-to-Tuberculosis disease diagnosis after completion of Tuberculosis preventive therapy among people living with HIV on Antiretroviral Therapy in Eastern Uganda: A retrospective cohort study

**DOI:** 10.64898/2026.04.08.26350451

**Authors:** Brian Atubu Esele, Bonniface Oryokot, Saadick Mugerwa Ssentongo, Muhammed Mulongo, Joyce Akanyo, Felix Bongomin

**Author notes:** Corresponding author; (BAE).

## Abstract

**Background:** Tuberculosis (TB) remains a leading cause of morbidity and mortality among people living with HIV (PLHIV), who face a 12-fold higher risk of active TB reactivation than HIV-negative individuals. TB preventive therapy (TPT) is an effective intervention, yet TB/HIV co-infection persists at 40–45%, raising questions about the durability of a single course of TPT. This study assessed the time from TPT completion to TB diagnosis and predictors of early TB reactivation.

**Methods:** We conducted a retrospective case-only cohort study using routine data from Uganda’s electronic medical record system, TB registers, and patient files at three TASO Centres of Excellence (Soroti, Mbale, Tororo). PLHIV on antiretroviral therapy (ART) diagnosed with TB after completing TPT between 2022–2024 were included. Participant characteristics and time to TB diagnosis were summarised descriptively; predictors of early TB were identified using logistic regression.

**Results:** Among 670 participants, most were female (464, 69.3%) with mean age 51.6 years (SD 14.5). Newly diagnosed TB accounted for 638 (95.2%), including bacteriologically confirmed pulmonary TB (535, 79.9%), clinically diagnosed TB (123, 18.4%), and extrapulmonary TB (12, 1.8%). Overall, 548 (82.8%) participants were virally suppressed, with most on Dolutegravir-based regimens (641, 95.7%). Early TB occurred in 144 (21.5%), with average time to diagnosis 2.6 years. Multivariable analysis showed care at TASO Soroti was protective (aOR = 0.104, p < 0.001), while clinically diagnosed TB (aOR = 1.91, p = 0.007), shorter ART duration (<5 years: aOR = 3.07, p = 0.001; 5–10 years: aOR = 1.74, p = 0.018), and viral suppression (aOR = 1.87, p = 0.014) increased odds of early TB.

**Conclusions:** TB can occur soon after TPT completion, with one in five PLHIV developing early disease particularly those with shorter ART duration despite viral suppression. Strengthening TB screening, continuous monitoring, and repeat TPT for high-risk groups may improve prevention.

## INTRODUCTION

Tuberculosis (TB) remains one of the leading causes of morbidity and mortality from infectious diseases globally [1]. According to the World Health Organization (WHO) Global Tuberculosis Report 2025, an estimated 10.7 million people developed TB in 2024, including approximately 5.8 million men, 3.7 million women, and 1.2 million children [2]. TB caused about 1.23 million deaths including 150,000 deaths among people living with HIV (PLHIV) worldwide [2]. Although global TB incidence has gradually declined in recent years, the disease continues to disproportionately affect low- and middle-income countries, with the highest burden occurring in South-East Asia (around 34%), Africa (approximately 25%), and the Western Pacific region (around 27%) [2]. Active TB disease manifests in several forms, including pulmonary and extrapulmonary disease. Pulmonary TB is particularly important for transmission because *Mycobacterium tuberculosis* (MTB) complex spreads through airborne particles expelled when infected individuals cough, sneeze, or speak [3].

Unlike active TB disease, latent tuberculosis infection (LTBI) is characterized by a persistent immune response to MTB antigens without clinical symptoms or signs of disease [4]. Although individuals with LTBI are asymptomatic and not infectious, they represent a substantial reservoir from which active TB disease may emerge [5]. Global LTBI prevalence declined from 30.66% in 1990 to 23.67% in 2019, but it remains a significant public health concern because of the large pool of individuals at risk of disease reactivation [6]. It is estimated that nearly one-quarter of the global population is infected with MTB, placing approximately 2 billion people at risk of developing active TB disease during their lifetime [2]. Among infected individuals, the lifetime risk of progression from latent infection to active disease is approximately 5–10%, with about half of this risk occurring within the first two years following infection [7,8]. Mathematical modelling studies suggest that this reservoir of latent infection will continue to generate substantial TB incidence in future decades despite improvements in TB control [9].

PLHIV are 12 times more likely to develop active TB disease compared with HIV-negative individuals [2]. HIV impairs cell-mediated immunity by targeting CD4-positive T lymphocytes, which play a critical role in controlling MTB infection. Consequently, PLHIV experience both increased susceptibility to new TB infection and higher rates of reactivation of latent infection [10]. Additional factors associated with TB activation include alcohol abuse, smoking, undernutrition, diabetes mellitus, and overcrowded living conditions [11–13].

To address this elevated risk among PLHIV, World Health Organization (WHO) has recommended tuberculosis preventive therapy (TPT) since 1998 as a key intervention to prevent progression from latent infection to active TB disease [14]. When optimally implemented, TPT has been shown to reduce TB incidence by 70–90% among PLHIV [15,16]. Consequently, TPT has been incorporated into comprehensive HIV care packages in many countries with high TB/HIV burden [17].

In Uganda, scale-up of TPT among PLHIV has been substantial in recent years. National program data indicate that TPT coverage increased from 0.6% in 2016 to approximately 88.8% in 2022 [18]. Despite this expansion, TB/HIV co-infection remains high, accounting for roughly 40% of all TB cases reported nationally [19]. One potential explanation is that most PLHIV receive TPT only once during their lifetime, after which they remain vulnerable to new TB exposures in high-transmission environments. The durability of protection following completion of TPT remains incompletely understood, particularly in settings with sustained TB transmission. Determining the time interval between TPT completion and subsequent TB diagnosis is therefore important for guiding future TB prevention strategies. This study aimed to determine the duration between TPT completion and TB diagnosis among PLHIV and to identify predictors of early TB occurrence after TPT completion. Findings from this study may inform policies regarding repeat or periodic TPT dosing to strengthen TB prevention among PLHIV in high-burden settings.

## METHODS

### Study design

We conducted a retrospective case-only cohort study of PLHIV on ART who developed TB after completing TPT between 1 January 2022 and 31 December 2024. The study used routinely collected clinical data obtained during routine HIV clinic visits. Data were extracted from the Uganda electronic medical record system (EMR version 3.4.1-alpha) and verified against TB treatment registers and patient medical files. Retrospective cohort designs are well suited for studying relatively rare outcomes using existing clinical datasets and allow efficient analysis of large patient cohorts.

This study was conducted and reported in accordance with the STROBE (Strengthening the Reporting of Observational Studies in Epidemiology) guidelines [20].

### Setting

The study was conducted at three Centres of Excellence (COEs) of The AIDS Support Organization (TASO) located in Soroti, Mbale, and Tororo districts in Eastern Uganda. These facilities provide comprehensive HIV and TB prevention, care, and treatment services in accordance with World Health Organization and Uganda Ministry of Health guidelines, including routine provision of TPT to PLHIV. Each COE serves an estimated catchment radius of approximately 75 km and collectively provides HIV care to about 22,000 PLHIV across roughly 28 districts in Eastern and North-Eastern Uganda. TASO facilities have reported high TPT completion rates of approximately 92%, substantially higher than the national average reported in public health facilities [21].

The three TASO Centers of Excellence (COEs) initiated TPT implementation in 2015, beginning with children living with HIV and subsequently expanding to the adult population. After a slow initial uptake, a national programmatic surge during July–September 2019 markedly accelerated implementation, enrolling more than 300,000 PLHIV in that quarter alone [22]. Eligibility screening was conducted using the WHO-recommended case finding form (CSF) to exclude active TB disease. Following initiation, PLHIV received both virtual and in-person adherence counselling to support completion of therapy. They also benefitted from regular assessments to identify potential toxicities for prompt interventions. Further, PLHIV initiating TPT also received pyridoxin to back stop potential side effects. On the other hand, individuals presenting with signs and symptoms suggestive of TB were further evaluated through sputum testing using WHO-recommended molecular rapid diagnostic tests (mWRDs), such as Xpert MTB/RIF or other nucleic acid amplification techniques. In addition, eligible PLHIV such as those with non-suppressed viral load underwent urine lipoarabinomannan (LAM) testing as a complementary diagnostic approach.

### Study population

The study included all PLHIV aged ≥5 years who were diagnosed with TB between 1 January 2022 and 31 December 2024 and had documented completion of TPT prior to TB diagnosis. Individuals without documented evidence of TPT completion were excluded.

### Inclusion and Exclusion criteria

**Inclusion criteria**: All PLHIV diagnosed with Tuberculosis between 1st January 2022 and 31st December 2024 and had completed TPT prior to diagnosis.

**Exclusion criteria**: All PLHIV diagnosed with Tuberculosis between 1st January 2022 and 31st December 2024 who received TPT but with no documented evidence of receipt or completion.

### Sampling approach

Total population sampling was used, whereby all eligible PLHIV meeting the inclusion criteria across the three TASO COEs during the study period were included in the analysis.

### Data Collection

Data were collected between 19^th^ March and 30^th^ June 2025. Lists of PLHIV diagnosed with TB during the study period were first extracted from TB treatment registers using ART clinic identification numbers. These identifiers were then used to retrieve corresponding clinical data from the Uganda EMR database. Missing information was verified using TPT registers and patient medical records. Extracted variables included demographic characteristics (age, sex, facility), ART initiation date, TPT initiation and completion dates, TB diagnosis date, WHO clinical stage, viral load, CD4 count, ART adherence level, and ART regimen.

### Operational definitions

The CD4 count, viral load (from plasma samples), WHO clinical stage, and adherence score used in analysis were the last documented values recorded before TB diagnosis. ART adherence was categorized according to the consolidated guidelines for the prevention and treatment of HIV and AIDs in Uganda [23]: good adherence (≥95%), fair adherence (85–94%), and poor adherence (<85%); Documented self-reported adherence feedback was used to capture data for this analysis. Viral load (from plasma samples) <1000 copies/mL was defined as suppressed, while ≥1000 copies/mL was considered non-suppressed. TB categories were defined as bacteriologically confirmed TB (if there was evidence for identification of mycobacteria from patients), clinically diagnosed TB (if patients were treated clinically without confirmed presence of mycobacteria), or extrapulmonary TB(for TB disease anatomical sites outside the lungs) according to national diagnostic criteria [24]. ART regimens were categorized as integrase strand transfer inhibitor (INSTI)–based, non-nucleoside reverse transcriptase inhibitor (NNRTI)–based, protease inhibitor (PI)–based, or complex/salvage regimens.

### Data Analysis

Data were cleaned and organized in Microsoft Excel before export to Stata for analysis. Descriptive statistics were used to summarize participant characteristics. Categorical variables were presented as frequencies and percentages, while continuous variables were summarized using means with standard deviations (SD) or medians with interquartile ranges (IQR).

Time to TB diagnosis after TPT completion was calculated as the interval between the TPT completion date and the TB diagnosis date. Early TB diagnosis was defined as TB occurring within 1.5 years after TPT completion, based on evidence suggesting a protective effect of TPT lasting approximately three years. Associations between potential predictors and early TB diagnosis were initially assessed using chi-square tests. Variables with p<0.2 and those supported by prior evidence were included in multivariable logistic regression models to identify independent predictors. Statistical significance was defined as p<0.05.

### Ethical Approval

Ethical approval was obtained from the TASO Research Ethics Committee (TASO-2025-326) on the 14^th^ March, 2025. A waiver of informed consent was granted because the study used routinely collected secondary data. Participant confidentiality was maintained by using unique clinic identification numbers rather than names, and all extracted datasets were anonymized prior to analysis. The study was conducted in accordance with STROBE reporting guidelines for observational studies.

## RESULTS

### Participant Characteristics

A total of 670 PLHIV who developed TB after completing TPT were included in the analysis. The majority were female (464, 69.3%), with a mean age of 51.6 years (SD 14.5) and a median age of 53 years (IQR 45–61). Most participants were aged >49 years (438, 65.4%), while children <15 years accounted for only 9 (1.3%) of the cohort. Participants were enrolled from three TASO Centres of Excellence: Tororo (395, 59.0%), Soroti (199, 29.7%), and Mbale (76, 11.3%) (Table 1).

**Table 1.** Sociodemographic and clinical characteristics of PLHIV diagnosed with tuberculosis after completion of tuberculosis preventive therapy (N = 670)

| Variable | n (%) |
| --- | --- |
| Health facility |  |
| TASO Mbale | 76 (11.3) |
| TASO Soroti | 199 (29.7) |
| TASO Tororo | 395 (59.0) |
| Age category |  |
| <15 years | 9 (1.3) |
| 15–49 years | 223 (33.3) |
| >49 years | 438 (65.4) |
| Median age (IQR) | 53 (45–61) |
| Mean age (SD) | 51.6 (14.5) |
| Sex |  |
| Female | 464 (69.3) |
| Male | 206 (30.8) |
| TB category |  |
| **Extrapulmonary TB** | 12 (1.8) |
| **Bacteriologically confirmed TB** | 535 (79.9) |
| **Clinically diagnosed TB** | 123 (18.4) |
| **Patient type** |  |
| **New TB case** | 638 (95.2) |
| **Relapse TB** | 32 (4.8) |
| **CD4 test done after TPT completion** |  |
| **No** | 237 (35.4) |
| **Yes** | 433 (64.6) |
| **CD4 category (n = 433)** |  |
| **≥200 cells/μL** | 374 (86.4) |
| **<200 cells/μL** | 59 (13.6) |
| **WHO clinical stage before TB diagnosis** |  |
| **Stage I** | 101 (15.1) |
| **Stage II** | 411 (61.3) |
| **Stage III** | 123 (18.4) |
| **Stage IV** | 35 (5.2) |
| **ART adherence** |  |
| **Fair (85–94%)** | 27 (4.0) |
| **Good (≥95%)** | 559 (83.4) |
| **Poor (<85%)** | 84 (12.5) |
| **Viral load suppression** |  |
| **Non-suppressed** | 122 (18.2) |
| **Suppressed** | 548 (82.8) |
| **Duration on ART** |  |
| **<5 years** | 63 (9.4) |
| **5–10 years** | 169 (25.2) |
| **>10 years** | 438 (63.4) |
| **ART regimen class** |  |
| **INSTI-based** | 641 (95.7) |
| **NNRTI-based** | 15 (2.2) |
| **PI-based** | 8 (1.2) |
| **Complex/salvage regimen** | 6 (0.9) |
| **ART regimen** |  |
| **TDF/3TC/DTG** | 619 (92.4) |
| **TDF/3TC/EFV** | 13 (1.9) |
| **TDF/3TC/ATV/r** | 2 (0.3) |
| **TDF/3TC/NVP** | 2 (0.3) |
| **ABC/3TC/DTG** | 15 (2.2) |
| **ABC/3TC/ATV/r** | 4 (0.6) |
| **AZT/3TC/DTG** | 7 (1.0) |
| **AZT/3TC/ATV/r** | 2 (0.3) |
| **TDF/3TC/DTG/DRV/RTV** | 5 (0.8) |
| **DTG/ETR/DRV/RTV** | 1 (0.2) |
| **Time to TB diagnosis after TPT completion** |  |
| **Mean years (SD)** | 2.62 (1.29) |
| **Median years (IQR)** | 2.53 (1.67–3.47) |
| **Early TB diagnosis (<1.5 years)** |  |
| **No** | 526 (78.5) |
| **Yes** | 144 (21.5) |

Abbreviations: TASO = The AIDS Support Organization; TB = tuberculosis; CD4 = cluster of differentiation 4; TPT = tuberculosis preventive therapy; WHO = World Health Organization; ART = antiretroviral therapy; INSTI = integrase strand transfer inhibitor; NNRTI = non-nucleoside reverse transcriptase inhibitor; PI = protease inhibitor; TDF = tenofovir disoproxil fumarate; 3TC = lamivudine; DTG = dolutegravir; EFV = efavirenz; ATV/r = atazanavir/ritonavir; NVP = nevirapine; ABC = abacavir; AZT = zidovudine; DRV/r = darunavir/ritonavir; ETR = etravirine; SD = standard deviation; IQR = interquartile range.

Most participants had been receiving antiretroviral therapy (ART) for more than 10 years (438, 63.4%), followed by 5–10 years (169, 25.2%) and <5 years (63, 9.4%). Most TB cases were newly diagnosed (638, 95.2%), while relapse TB accounted for 32 (4.8%). Bacteriologically confirmed pulmonary TB constituted 535 (79.9%) of cases, clinically diagnosed pulmonary TB 123 (18.4%), and extrapulmonary TB 12 (1.8%) (Table 1).

Among the 433 participants (64.6%) with available CD4 results, most had CD4 counts ≥200 cells/µL (374, 86.4%), while 59 (13.6%) had CD4 counts <200 cells/µL. Most participants were classified as WHO clinical stage II (411, 61.3%), followed by stage III (123, 18.4%), stage I (101, 15.1%), and stage IV (35, 5.2%). Viral load suppression (<1000 copies/mL) was observed in 548 participants (82.8%). Most participants were receiving integrase strand transfer inhibitor (INSTI), based ART regimens (641, 95.7%) (Table 1).

### Time to TB diagnosis after TPT completion

The mean duration between completion of TPT and TB diagnosis was 2.62 years (SD 1.29), while the median time was 2.53 years (IQR 1.67–3.47) (Table 1). Overall, 144 participants (21.5%) developed TB within 1.5 years after completing TPT and were classified as having early TB, while 526 (78.5%) developed TB after this period.

### Factors associated with early TB diagnosis

Bivariate analysis identified several factors associated with early TB diagnosis after TPT completion, including health facility, TB diagnostic category, CD4 testing status, ART adherence, duration on ART, and viral load suppression (Table 2).

**Table 2.** Bivariate analysis of factors associated with early Tuberculosis diagnosis after tuberculosis preventive therapy completion (N = 670)

| Variable | No early TB n (%) | Early TB n (%) | p-value |
| --- | --- | --- | --- |
| **Health facility** |  |  | 0.001 |
| TASO Mbale | 53 (69.7) | 23 (30.3) |  |
| TASO Soroti | 191 (96.0) | 8 (4.0) |  |
| TASO Tororo | 282 (71.4) | 113 (28.6) |  |
| **Age category** |  |  | 0.077 |
| <15 years | 6 (66.7) | 3 (33.3) |  |
| 15–49 years | 165 (74.0) | 58 (26.0) |  |
| >49 years | 355 (81.0) | 83 (19.0) |  |
| **Sex** |  |  | 0.955 |
| Female | 364 (78.4) | 100 (21.6) |  |
| Male | 162 (78.6) | 44 (21.4) |  |
| **TB category** |  |  | 0.001 |
| Extrapulmonary TB | 9 (75.0) | 3 (25.0) |  |
| Bacteriologically confirmed TB | 436 (81.5) | 99 (18.5) |  |
| Clinically diagnosed TB | 81 (65.9) | 42 (34.2) |  |
| **Patient type** |  |  | 0.408 |
| New TB | 499 (78.2) | 139 (21.8) |  |
| Relapse TB | 27 (84.4) | 5 (15.6) |  |
| **CD4 done** |  |  | 0.001 |
| No | 166 (70.0) | 71 (30.0) |  |
| Yes | 360 (83.1) | 73 (16.9) |  |
| **ART adherence** |  |  | 0.010 |
| Fair | 24 (88.9) | 3 (11.1) |  |
| Good | 446 (79.9) | 113 (20.2) |  |
| Poor | 56 (66.7) | 28 (33.3) |  |
| **Duration on ART** |  |  | 0.001 |
| <5 years | 33 (52.4) | 30 (47.6) |  |
| 5–10 years | 124 (73.4) | 45 (26.6) |  |
| >10 years | 369 (84.3) | 69 (15.7) |  |
| **Viral load suppression** |  |  | 0.009 |
| Non-suppressed | 85 (69.7) | 37 (30.3) |  |
| Suppressed | 441 (80.5) | 107 (19.5) |  |
| **ART regimen class** |  |  | 0.633 |
| INSTI-based | 504 (78.6) | 137 (21.4) |  |
| NNRTI-based | 10 (66.7) | 5 (33.3) |  |
| PI-based | 7 (87.5) | 1 (12.5) |  |
| Complex regimen | 5 (83.3) | 1 (16.7) |  |

Abbreviations: TASO = The AIDS Support Organization; TB = tuberculosis; CD4 = cluster of differentiation 4; ART = antiretroviral therapy; INSTI = integrase strand transfer inhibitor; NNRTI = non-nucleoside reverse transcriptase inhibitor; PI = protease inhibitor.

**Table 3.** Multivariable predictors of early tuberculosis diagnosis after tuberculosis preventive therapy completion.

| Variable | Crude OR (95% CI) | p-value | Adjusted OR (95% CI) | p-value |
| --- | --- | --- | --- | --- |
| **Health facility** |  |  |  |  |
| TASO Mbale | Reference |  | Reference |  |
| TASO Soroti | 0.09 (0.04–0.23) | <0.001 | 0.10 (0.04–0.23) | <0.001 |
| TASO Tororo | 0.92 (0.54–1.57) | 0.771 | 0.92 (0.52–1.62) | 0.775 |
| **TB category** |  |  |  |  |
| Bacteriologically confirmed | Reference |  | Reference |  |
| Clinically diagnosed | 2.28 (1.48–3.51) | 0.001 | 1.91 (1.19–3.06) | 0.007 |
| Extrapulmonary TB | 1.46 (0.39–5.52) | 0.570 | 1.14 (0.29–4.42) | 0.842 |
| **CD4 done** |  |  |  |  |
| No | Reference |  | Reference |  |
| Yes | 0.47 (0.32–0.68) | 0.001 | 0.99 (0.64–1.57) | 0.997 |
| **Duration on ART** |  |  |  |  |
| >10 years | Reference |  | Reference |  |
| 5–10 years | 1.94 (1.26–2.97) | 0.002 | 1.74 (1.09–2.75) | 0.018 |
| <5 years | 4.86 (2.78–8.48) | 0.001 | 3.07 (1.61–5.88) | 0.001 |
| **Viral load suppression** |  |  |  |  |
| Non-suppressed | Reference |  | Reference |  |
| Suppressed | 1.79 (1.15–2.78) | 0.009 | 1.87 (1.13–3.08) | 0.014 |

Abbreviations: TASO = The AIDS Support Organization; TB = tuberculosis; CD4 = cluster of differentiation 4; ART = antiretroviral therapy.

In multivariable analysis, the odds of early TB varied significantly by health facility. Patients enrolled at TASO Soroti had markedly lower odds of early TB compared to TASO Mbale (aOR = 0.10, 95% CI: 0.04– 0.23, p<0.001), while no difference was observed for TASO Tororo (aOR = 0.92, p=0.775). Clinically diagnosed TB cases were more likely to present as early TB compared to bacteriologically confirmed cases (aOR = 1.91, 95% CI: 1.19–3.06, p=0.007), whereas extrapulmonary TB showed no significant association (aOR = 1.14, p=0.842). Having a CD4 test done was protective in crude analysis but lost significance after adjustment (aOR = 0.99, p=0.997). Duration on ART was strongly associated with early TB: individuals on ART for less than five years had threefold higher odds (aOR = 3.07, 95% CI: 1.61– 5.88, p=0.001), and those on ART for 5–10 years had nearly twofold higher odds (aOR = 1.74, 95% CI: 1.09–2.75, p=0.018), compared to those on ART for more than 10 years. Interestingly, viral load suppression was associated with increased odds of early TB (aOR = 1.87, 95% CI: 1.13–3.08, p=0.014).

## DISCUSSION

This study examined the timing of TB diagnosis following completion of TPT among 670 PLHIV receiving care at three HIV treatment centres in Eastern Uganda. The mean duration from TPT completion to TB diagnosis was 2.6 years (approximately 31 months). Early TB diagnosis, defined as TB occurring within 1.5 years after TPT completion, was associated with the health facility where care was received, clinically diagnosed TB, shorter duration on ART, and viral load suppression.

There is limited evidence describing the duration of protection following completion of TPT. In this study, TB occurred on average 2.6 years after TPT completion, suggesting that the protective effect of a single TPT course may diminish over time. This finding is broadly consistent with evidence from Ethiopia indicating that the protective benefit of TPT is strongest within the first three years following therapy [25]. A previous Ugandan study reported a shorter duration of 18.5 months; however, that study measured time from TPT initiation rather than completion and involved a much smaller sample size [26].

Declining protection over time has also been reported in other cohorts of PLHIV receiving isoniazid preventive therapy [25,27]. These observations support growing discussion around repeat or extended TPT strategies in high-burden settings. Uganda currently recommends a single course of TPT for PLHIV, yet TB/HIV co-infection remains high, with approximately 40–43% of TB patients living with HIV [19,28]. Our findings therefore reinforce the need to re-evaluate long-term TB prevention strategies for PLHIV in settings with sustained community transmission.

Health facility was an important predictor of early TB diagnosis, with participants receiving care at TASO Soroti having significantly lower odds of early TB compared with those receiving care at TASO Mbale. This finding suggests potential inter-facility differences in TB prevention and clinical care. Variability between health facilities may arise from differences in diagnostic capacity, screening practices, infection prevention and control measures, and availability of trained healthcare personnel [29–31].

In Uganda, health facilities with sustained funding and strong programmatic support from partners such as USAID and the Center for Disease Control and Prevention have been shown to perform better in TB case detection and prevention activities [32]. Programmatic strengths such as improved follow-up systems, adherence counselling, and infection control practices may partly explain the protective effect observed at TASO Soroti. Additionally, regional differences in TB transmission may contribute to these findings, as TB/HIV burdens vary across districts in Eastern Uganda [33].

Shorter duration on ART was strongly associated with early TB diagnosis. Participants on ART for less than five years had over threefold higher odds of early TB compared with those on ART for more than ten years. Similar patterns have been observed in multiple settings. For example, a Tanzanian cohort study reported substantially higher TB incidence during the early months following ART initiation, with risk declining as ART duration increased [34]. Studies from Europe have also documented increased TB risk during the first 3–6 months of ART, particularly among individuals with severe immunosuppression [35–37].

One explanation for this phenomenon is immune reconstitution inflammatory syndrome (IRIS), which may occur soon after ART initiation as immune function begins to recover [38,39]. Immune reconstitution can unmask previously undiagnosed TB or trigger inflammatory responses to latent infections. Over time, sustained ART leads to improved immune recovery and increasing CD4 cell counts, which are inversely associated with TB risk [36,40]. Long-term ART is therefore a critical protective factor against TB among PLHIV.

Clinically diagnosed TB was associated with higher odds of early TB diagnosis compared with bacteriologically confirmed TB. Although most cases in this study were bacteriologically confirmed, which reflects strong diagnostic capacity within the participating facilities, clinically diagnosed TB may occur more frequently in the early stages of disease when bacterial load is low and laboratory confirmation is difficult [41,42]. In such situations, clinicians often rely on symptoms and radiological findings to initiate treatment promptly [43,44].

Clinical diagnosis may also be more common among patient groups such as children or individuals unable to produce sputum, where bacteriological confirmation is challenging [45]. However, reliance on clinical diagnosis carries the risk of overdiagnosis and false positive TB diagnoses, which is why many guidelines emphasize bacteriological confirmation whenever possible [3,46].

Unexpectedly, viral load suppression was associated with higher odds of early TB diagnosis. Viral suppression is generally associated with improved immune recovery and reduced risk of opportunistic infections [47]. Indeed, ART and viral suppression have been shown to reduce TB risk by up to 68% among PLHIV [25,27,48].

Several explanations may account for this finding. First, PLHIV with viral suppression often remain engaged in care and attend clinic visits regularly, increasing the likelihood of routine TB screening and early detection [49,50]. Second, persistent community-level TB transmission in high-burden settings such as Uganda may result in repeated exposure even among individuals with good HIV control [51,52]. Third, non-HIV-related risk factors, including poverty, overcrowding, malnutrition, diabetes, smoking, and alcohol use, may independently increase TB risk despite viral suppression [11,53]. Finally, delayed immune recovery may occur in some individuals despite virologic suppression, particularly among those initiating ART with advanced immunosuppression [54–56].

### Strengths and limitations

This study used routinely collected clinical data from large HIV treatment cohorts, providing findings that reflect real-world programmatic conditions. The use of total population sampling minimized selection bias and enabled inclusion of a relatively large sample of PLHIV diagnosed with TB after TPT completion.

However, several limitations should be considered. First, the use of secondary data limited the availability of important variables such as smoking status, alcohol use, diabetes, and household TB exposure, which are known risk factors for TB. Second, data quality depended on routine clinical documentation, which may introduce information bias [57]. To mitigate this limitation, data were triangulated across multiple sources including TB registers, electronic medical records, and patient files.

## Conclusions

Most TB diagnoses among PLHIV in this cohort occurred within approximately 30 months after completion of TPT, suggesting that the protective effect of a single TPT course may decline over time in high-burden settings. Importantly, TB occurred even among individuals with viral suppression, particularly among those with shorter durations on ART. These findings highlight the need for sustained TB screening and prevention strategies among PLHIV beyond initial TPT completion. In high TB transmission settings, consideration should be given to repeat or extended TPT strategies, particularly for individuals at increased risk of early TB after completing preventive therapy.

## Data Availability

All relevant data underlying the findings of this study are fully available within the manuscript and its Supporting Information files. Additional data may be obtained from the corresponding author upon reasonable request.

## Acknowledgment

The author(s) thank Georgios Kitsaras and Dr. Welike Emmanuel for their guidance, acknowledge the great role of the data collectors led by Ogwang Calvin, TASO and staff and appreciate the support of their families throughout this research.

## Availability of data and materials

All relevant data are within the manuscript and its supporting information files. Data are available upon reasonable request from the first author.

## Consent for publication

Not applicable.

## AUTHOR CONTRIBUTIONS

**Conceptualization**: Brian Atubu Esele, Bonniface Oryokot, Felix Bongomin.

**Data curation**: Brian Atubu Esele, Bonniface Oryokot, Saadick Mugerwa Ssentongo, Muhammed Mulongo, Joyce Akanyo.

**Formal analysis**: Brian Atubu Esele, Bonniface Oryokot, Saadick Mugerwa Ssentongo, Felix Bongomin.

**Investigation:** Brian Atubu Esele, Bonniface Oryokot, Saadick Mugerwa Ssentongo, Muhammed Mulongo, Joyce Akanyo.

**Methodology**: Brian Atubu Esele, Bonniface Oryokot, Saadick Mugerwa Ssentongo, Muhammed Mulongo, Joyce Akanyo, Felix Bongomin.

**Project administration**: Brian Atubu Esele, Bonniface Oryokot, Muhammed Mulongo.

**Software**: Brian Atubu Esele.

**Supervision**: Felix Bongomin.

**Validation**: Brian Atubu Esele, Joyce Akanyo, Felix Bongomin, Bonniface Oryokot

**Visualization**: Brian Atubu Esele, Bonniface Oryokot, Felix Bongomin.

**Writing** – **original draft:** Brian Atubu Esele, Akanyo Joyce and Felix Bongomin

**Writing – review & editing**: Brian Atubu Esele, Bonniface Oryokot, Saadick Mugerwa Ssentongo, Muhammed Mulongo, Joyce Akanyo, Felix Bongomin.

